# Paediatric escalation area and corridor care in the Emergency Department, a secondary analysis of the UNCORKED study

**DOI:** 10.64898/2026.09.02.26362046

**Authors:** Chidi Anakebe, Daniel R Owens, Damian Roland, Fraser Birse, Ryan McHenry, Ben Clarke, Tom Roberts, Trainee Emergency Research Network UNCORKED study group

## Abstract

**Objective:** To describe the use of, and outcomes following, Emergency Department (ED) escalation area care among children.

**Design:** Two-stage prospective multicentre observational study. Stage one was a cross-sectional snapshot study reporting the point prevalence of escalation area care; stage two was an observational cohort study of eligible participants present in the EDs during snapshots.

**Setting:** 5 snapshots of 96 type 1 UK EDs. 83/96 (86.5%) of sites were mixed EDs and 13 sites were paediatric-only EDs.

**Patients:** Patient level data was collected on those who experienced escalation area care and/or were admitted.

**Outcomes:** The primary outcome was the proportion of patients cared for in escalation areas in paediatric-only EDs. Secondary outcomes were demographics, length of stay (LOS), 28-day mortality, type of escalation area experienced and time in escalation areas in all ED types; stratified by escalation area experience.

**Results:** 6.6% (n=75/1132) of patients in paediatric-only EDs were receiving escalation area care during the study snapshots, the majority of this occurred in a minority of departments. Across all ED types, 15.4% (n=104/675) of admitted children experienced escalation area care during their ED stay. 124/244 (50.8%) of those cared for in escalation areas were cared for in a non-clinical area such as a corridor. 9.6% (n=8/83) of mixed EDs reported caring for adults in paediatric areas. Three children died within 28 days; all admitted to hospital without experiencing escalation area care. Hospital LOS was 2.00 days (1.00, 3.00) for those in escalation areas and 2.00 days (2.00, 4.00) for those not cared for in escalation areas.

**Conclusions:** Escalation area care is a common and systemic feature of paediatric emergency care in the UK which requires urgent action to ensure children are cared for in appropriate environments.

## Introduction

There is rising concern in the United Kingdom (UK) about the use of Emergency Department (ED) escalation area care, also known as ‘corridor’ or ‘hallway’ care, among This concern comes from clinicians, policy makers, hospital management and most importantly patients and their families.^1,2,3^

Escalation area care can be described as patient care that is delivered in areas of the ED not designed for that purpose. One of the key challenges has been a lack of an accepted definition for “escalation area care” and NHS England did not define it until March 2026.^4^ These areas may be non-clinical areas such as a corridor or repurposed clinical areas such as outpatient clinics or ‘doubling-up’ patients in a cubicle. Escalation area use impacts patient dignity and there is a concern that it poses a significant patient safety risk. ^5,6^ To date, the focus of research has been on the impact of escalation area care on adult patients, but little is known about the scale or impact of escalation area care on children attending the ED.

In the UK, Royal College of Emergency Medicine (RCEM) guidance states that children should never be cared for in escalation areas.^7^ Furthermore, Royal College of Paediatrics and Child Health (RCPCH) guidance states that children in the ED should have separate waiting and treatment areas from adults to ensure that children are managed and safeguarded in a child-friendly environment.^8^ It is therefore important to understand if children are being cared for in escalation areas and if system-wide pressures are resulting in adults being cared for in paediatric areas in mixed adult and paediatric EDs.

The ‘Understanding escalation area and corridor care in UK emergency departments’ (UNCORKED) study was a UK-wide multi-centre study assessing the impact of escalation area use on adult and paediatric patients. Results from the UNCORKED study published in 2026 highlighted that at any one time, one in five adult patients are receiving escalation area care and one in three adult patients admitted to hospital spent time in ED corridors prior to admission. Furthermore, up to 26% of EDs had no resuscitation cubicles available.^9^ It is unknown if this situation is replicated for paediatric patients. This analysis aims to describe the use of, and outcomes following, ED escalation area care among children (<16 years).

## Methods

### Study design and setting

A two stage prospective multicentre observational study led by the Royal College of Emergency Medicine’s (RCEM) Trainee Emergency Research Network (TERN). Stage one was a cross-sectional snapshot study reporting the point prevalence of escalation area care; stage two was an observational cohort study that recruited eligible participants present in the ED during the snapshots.

All UK Type 1 EDs (those providing consultant-led, 24-hour services with full resuscitation facilities) were eligible to participate. Site engagement was achieved by email dissemination to the TERN mailing list and through the National Institute of Health Research (NIHR) regional research delivery networks (RDNs). In this unplanned sub-analysis, data was only included from sites that saw paediatric patients (<16 years), i.e. mixed (adult and paediatric) and paediatric-only EDs. Although site-level data was collected from 13 EDs in Scotland, no patient-level data was collected from Scottish EDs, due to differing regulatory approval of recruitment via waived consent.

Eligible patients were recruited at five predetermined snapshots over 14 days, chosen to give a representative spread of ED activity based on published attendance data:^9^

1. 12:00, 03/03/2025 (Monday)
2. 07:00, 06/03/2025 (Thursday)
3. 16:00, 08/03/2025 (Saturday)
4. 19:00, 10/03/2025 (Monday)
5. 23:59, 12/03/2025 (Wednesday)

### Participants

Trained clinical research nurses and Emergency Medicine clinicians prospectively identified all patients present in the ED during the snapshots. The total number of patients present in the EDs at each snapshot was recorded as a denominator. They subsequently screened each patient’s ED attendance to determine whether they were: admitted to hospital having experienced escalation area care; admitted without having experienced escalation area care; discharged from the ED having experienced escalation area care; discharged from the ED without experiencing escalation area care.

All patients, except those who were discharged without experiencing escalation area care, were recruited to the study. Demographics and outcome data of those discharged having not experienced escalation area care were not collected to ensure feasibility. There were no additional exclusion criteria. Patients were recruited via waived consent. The patient level data for adult patients (≥16 years of age) will be reported separately.

There is no universally agreed definition for an escalation area. The following definition was provided to sites: ‘any area not routinely used unless the capacity of the usual ED geographical footprint is exceeded.’ Sites were asked to assign each escalation area to one of the following categories:

- Ambulance queueing to offload for >15 minutes.
- Repurposed clinical area.
- Non-clinical area (e.g. hospital corridor).
- Doubled-up cubicle.

Patients in the waiting room were considered to be in a non-clinical escalation area if there was objective evidence that they would be in a standard ED cubicle if one were available. For example, a patient receiving intravenous medication, and/or awaiting an inpatient bed whilst in a waiting room. Other than queuing ambulances and pre-hospital cohort areas, only patients in escalation areas under the care of the ED team were included. The central study team liaised with sites to identify relevant areas within their EDs.

### Outcomes

The primary outcome was the proportion of all paediatric patients present in participating EDs that were cared for in an escalation area at any point during their ED stay. This is reported solely for paediatric-only EDs as adult patients would be included in the denominator for mixed departments whilst being excluded from the numerator.

Secondary outcomes were descriptive analyses of children in all ED types, stratified by escalation area experience, of patient demographics, hospital LOS (in days), 28-day all-cause mortality, type of escalation area experienced, ED LOS, and time spent in escalation areas (both reported in hours).

### Data Collection

Local study teams prospectively identified all patients present in the ED during snapshots.

They subsequently used electronic health records (EHR), department management systems and in-department observation to determine patients’ disposition from the ED, whether they experienced escalation area care and the duration of time spent in escalation areas. Patient demographics and diagnosis were collected from patient records, and IMD from the registered postcode using a nation-specific tool. Hospital LOS was collected from hospital systems and discharge letters. At 28-days, hospital and GP records were examined for evidence of patient death. Data was entered directly into REDCap.

### Statistical analysis plan

The characteristics of participating sites are reported as the number and percentage of total sites relevant to this sub-analysis which were sites recruiting paediatric patients. Missing data is indicated in the results, and sites reporting inconsistent numbers between the total number in escalation areas and those in specific sub-areas were considered missing.

Inconsistencies in the description of ED characteristics (such as trauma status) between snapshots were resolved by acceptance of the most commonly reported characteristics.

As no relevant denominator of the total number of paediatric patients was available for mixed EDs, it is not possible to report a meaningful number, or a proportion, of paediatric patients experiencing escalation area care for these departments. Paediatric-only EDs were assessed as a subgroup of relevance, and one where the total numbers in each ED at each snapshot allowed presentation of results with a meaningful denominator. For this subgroup, the total number and proportion of patients in escalation areas, and admitted, was reported.

### Ethical approval and study conduct

Ethical approval was gained from a Brighton and Sussex research ethics committee (ref: 24/LO/0837). Regulatory approval was obtained from the Health Regulation Authority (HRA) and Health and Care Research Wales (HCRW). The study was conducted in accordance with the UK Policy Framework for Health and Social Care Research and other applicable guidance.

### PPI and Stakeholder engagement

The RCEM Lay advisory Group were involved during the study design, commenting specifically on the importance of the research questions and acceptability of the methodology, including the model of consent.

## Results

### Site characteristics

96 sites submitted paediatric patient data for at least one snapshot (**Error! Reference source not found.**). 83/96 (86.5%) sites were mixed EDs and 13 (13.5%) sites were paediatric only EDs. Patients were recruited from 88 sites in England, 6 in Wales and 2 in Northern Ireland. One site was unable to recruit individual patients due to issues with their EHR and was excluded, two sites previously reporting aggregated adult and paediatric ED data reported their paediatric population separately for this analysis. 61/96 (63.5%) were Trauma Units, 22/96 (22.9%) were Local Emergency Hospitals and 13/96 (13.5%) were Major Trauma Centres.

### Escalation area care in paediatric-only emergency departments by snapshot

Of the 96 sites recruiting paediatric patients, 13 were paediatric-only EDs, with between 11 and 12 collecting data at each snapshot. Between 16.7% (n=2/12) and 45.5% (n=5/11) reported escalation area use depending on the snapshot. Across all snapshots, 1,132 patients were present in paediatric-only EDs reporting occupancy, of which 6.6% (n=75/1,132) were seen to be in an escalation area. Of the 11 paediatric only EDs reporting resuscitation cubicle availability, between 9.1% (n=1/12) and 27.3% (n=3/11) reported having no available resuscitation cubicle during the snapshots. Table 2 demonstrates the number and proportion of patients experiencing escalation area care, and admissions, in paediatric only EDs by snapshot.

**Table 1.** Characteristics of participating EDs (N=96).

| Characteristic |  |  |
| --- | --- | --- |
| Region |  |  |
| East of England | n (%) | 9 (9.4) |
| London |  | 14 (14.6) |
| Midlands |  | 13 (13.5) |
| North East and Yorkshire |  | 10 (10.4) |
| North West |  | 11 (11.5) |
| South East |  | 18 (18.8) |
| South West |  | 13 (13.5) |
| Northern Ireland |  | 2 (2.1) |
| Wales |  | 6 (6.2) |
| Hospital Trauma-Receiving Status |  |  |
| Local Emergency Hospital | n (%) | 22 (22.9) |
| Major Trauma Centre |  | 13 (13.5) |
| Trauma Unit |  | 61 (63.5) |
| Hospital Age-Receiving Status |  |  |
| Mixed Adult/Paediatric |  | 83 (86.5) |
| Paediatric-only |  | 13 (13.5) |
| Patients Recruited Per Site |  |  |
| Admitted and in Escalation | Median (IQR) | 2 (1-4) |
| Admitted and not in Escalation |  | 5 (3-8) |
| Not Admitted and in Escalation |  | 2 (1-3.75) |

**Table 2.**
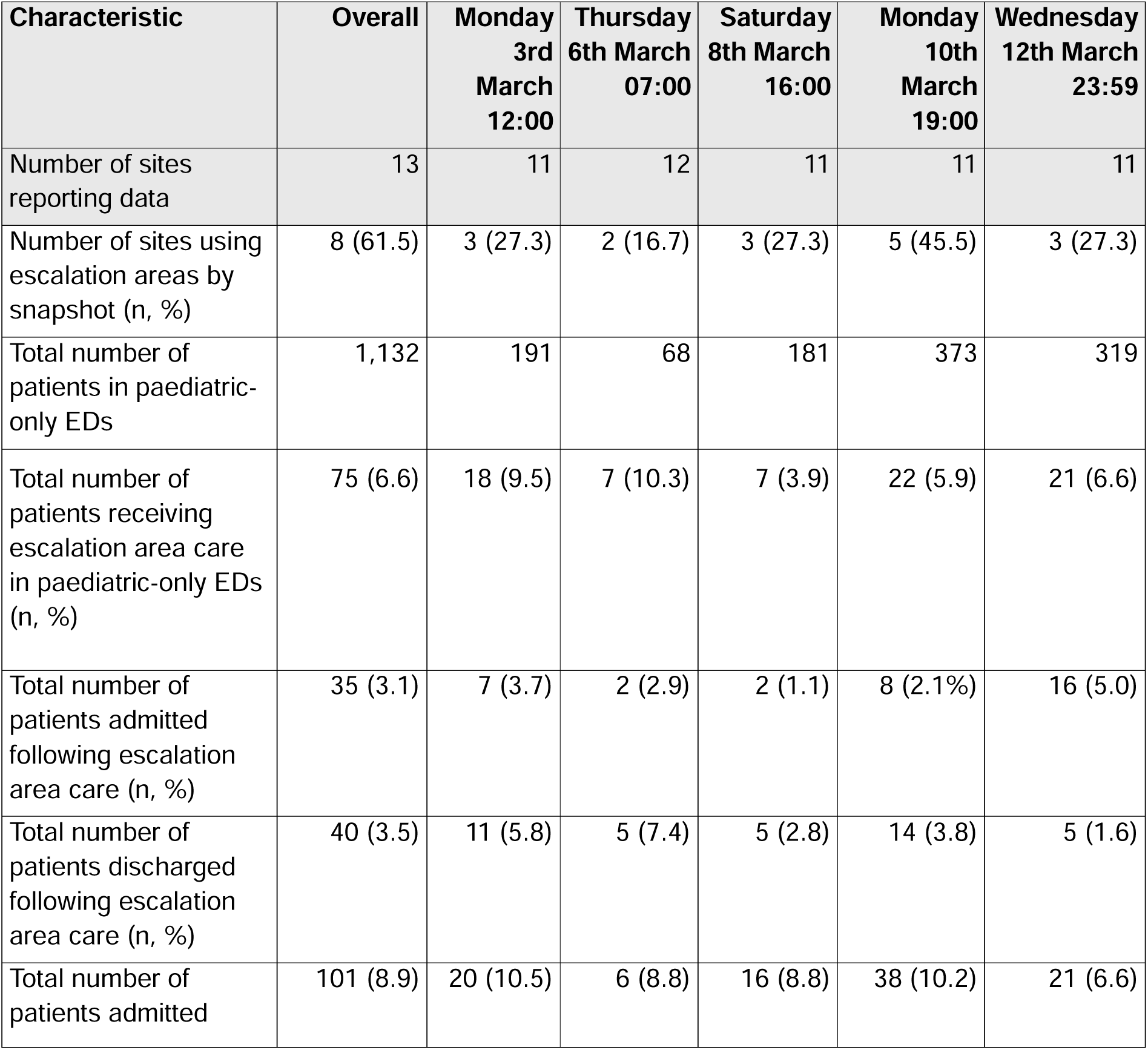
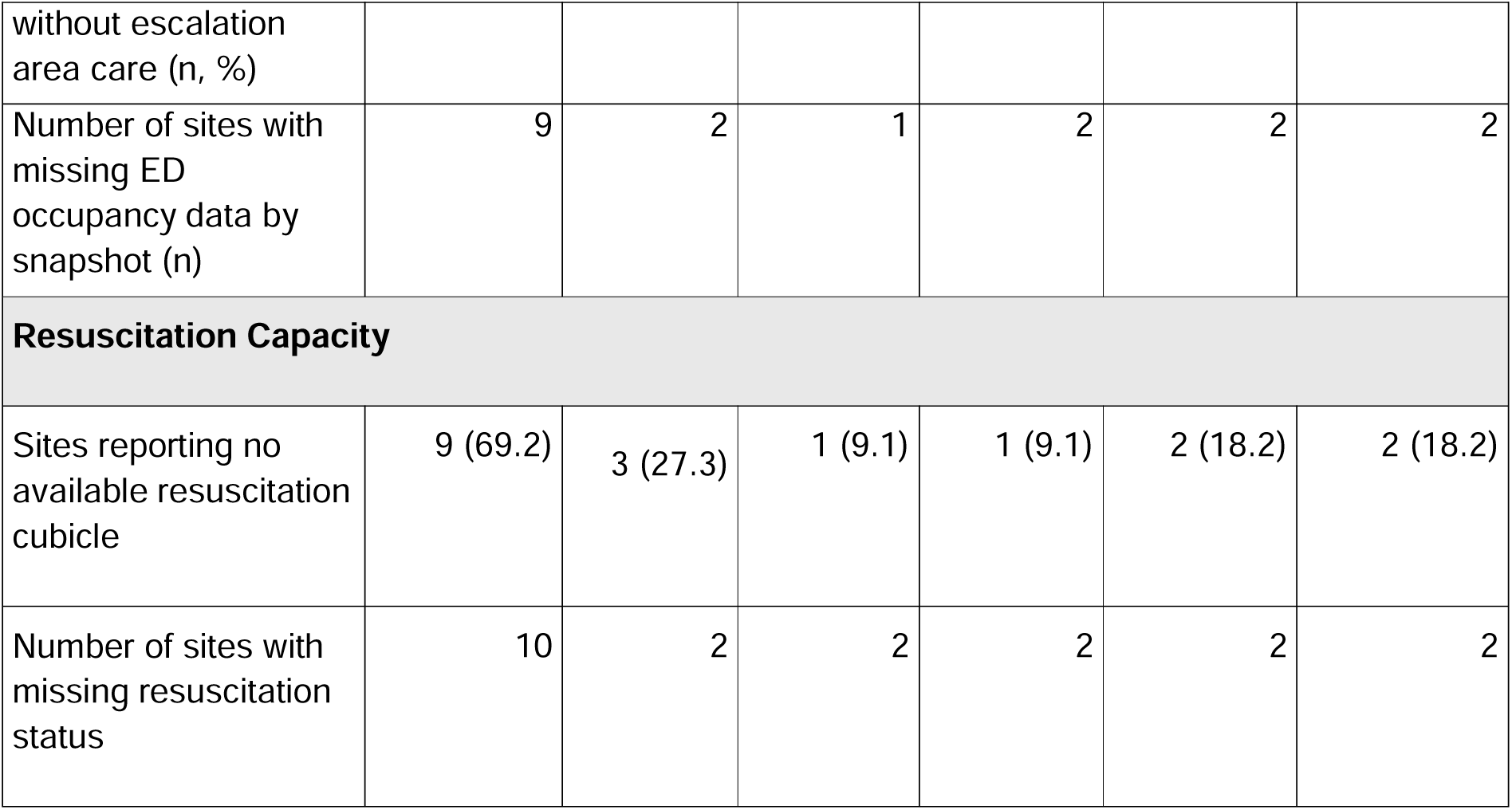
Number and proportion of paediatric-only sites using escalation area care, and of patients in escalation area care, or admitted, overall and for each snapshot. Note that 11-12 of the 13 sites reported data for each snapshot.

### Patients recruited from mixed and paediatric-only emergency departments

From all 96 sites (mixed and paediatric-only), a total of 815 eligible patients (<16 years of age, and either experiencing escalation area care or admitted) were recruited across the 5 snapshots (Table 3). 70.1% (n=571/815) of these were children admitted without experiencing escalation area care, 17.2% (n=140/815) were treated in an escalation area and subsequently discharged and 12.8% (n= 104/815) were cared for in escalation area and subsequently admitted. 9.6% (n=8/83) of mixed EDs reported the use of paediatric areas to care for adult patients during at least one snapshot.

**Table 3.** Demographics of children captured during snapshots.

| Characteristic | Statistic | Overall<br>N = 815 | Admitted<br>and in<br>Escalation N<br>= 104 | Admitted and<br>not in<br>Escalation N<br>= 571 | Not Admitted<br>and in<br>Escalation N<br>= 140 |
| --- | --- | --- | --- | --- | --- |
| **Age** | Median<br>(Q1, Q3) | 4 (1, 11) | 2 (1, 9) | 4 (1, 9) | 8 (2, 13) |
| **Sex** |  |  |  |  |  |
| Male | n (%) | 433<br>(53.4) | 50 (48.5) | 307 (54.0) | 76 (54.3) |
| Female |  | 378<br>(46.6) | 53 (51.5) | 261 (46.0) | 64 (45.7) |
| Unknown | n | 4 | 1 | 3 | 0 |
| **Ethnicity** |  |  |  |  |  |
| Asian | n (%) | 88<br>(13.0) | 10 (11.0) | 64 (13.6) | 14 (12.1) |
| Black |  | 39 (5.7) | 4 (4.4) | 26 (5.5) | 9 (7.8) |
| Mixed |  | 36(5.3) | 6 (6.6) | 22 (4.7) | 8 (6.9) |
| Other |  | 21(3.1) | 1 (1.1) | 16 (3.4) | 4 (3.4) |
| White |  | 495.0<br>(72.9) | 70 (76.9) | 344 (72.9) | 81 (69.8) |
| Unknown | n | 136 | 13 | 99 | 24 |
| **Index of**<br>**Multiple**<br>**Deprivation**<br>**Decile** | Median<br>(Q1, Q3) | 5 (3, 7) | 6. (3, 8) | 5 (2, 7) | 4.5 (3, 7.) |
| Unknown | n | 7 | 1 | 6 | 0 |

### Demographics

The median age of all children recruited was 4 years (IQR 1-11), see Table 3. The median age of children that were admitted having experienced escalation area care was 2 (IQR 1-9) compared to 4 (IQR 1-9) for those who were admitted without experiencing escalation area care, and 8 (IQR 2-13) for those cared for in escalation areas and discharged. 53% (n=433/815) of recruited children were male and 47% (n=378/815) were female. 73% (n=495/815) of children were identified as of white ethnicity, 13% (n=88/815) were Asian, 6% (n=39/815) Black ethnicity and 5% (n=36/815) as mixed ethnicity.

The median Index of Multiple deprivation (IMD) decile of all children captured during the study was 5 (IQR 3-7). The median IMD of children that were admitted without experiencing escalation area care was 5 (IQR 2-7) and of children that were admitted having experienced escalation area care was 6 (IQR 3-8).

### ED LOS, type of escalation area experienced, time spent in escalation areas

Median ED LOS of all recruited patients was 5.4 hours (IQR 3.5-81). For those admitted having experienced escalation area care it was 7.1 hours (IQR 4.5-10.9), for those admitted without experiencing escalation area care it was 5.5 hours (IQR 3.6-8.0) and for those who experienced escalation area care and when then discharged it was 4.0 hours (IQR 2.8-6.5). For those spending any time in an escalation area, the median LOS in those areas was 2.73 hours (IQR 0.98-5.04 hours) for those admitted, and 1.84 hours (IQR 0.89-3.24 hours) for those subsequently discharged. See Table 4.

**Table 4.** Outcomes of children stratified by escalation area placement and admission status.

| **Characteristic** | **Statistic** | **Overall**<br>**N = 815** | **Admitted**<br>**and in**<br>**Escalation**<br>**N = 104** | **Admitted**<br>**and not in**<br>**Escalation N**<br>**= 571** | **Not**<br>**Admitted**<br>**and in**<br>**Escalation N**<br>**= 140** |
| --- | --- | --- | --- | --- | --- |
| ED Length of Stay (Hours) | Median (Q1, Q3) | 5.4 (3.5, 8.1) | 7.1 (4.5, 10.8) | 5.5 (3.6, 8.0) | 4.0 (2.8, 6.5) |
| Total Duration in Escalation Areas (Hours) | Median (Q1, Q3) | 0.00 (0.00, 0.68) | 2.73 (0.98, 5.04) | 0.00 (0.00, 0.00) | 1.84 (0.89, 3.24) |
| Any Non-Clinical Escalation Area/'Corridor' Care | n (%) | 124.0 (15.2) | 55.0 (52.9) | 0.0 (0.0) | 69.0 (49.3) |
| 28 Day Mortality | n (%) | 3.0 (0.4) | 0.0 (0.0) | 3.0 (0.5) | 0.0 (0.0) |
| Hospital Length of Stay (Days) | Median (Q1, Q3) | 2.00 (1.00, 3.00) | 2.00 (1.00, 3.00) | 2.00 (2.00, 4.00) | 1.00 (1.00, 1.50) |
| Unknown | n | 14 | 1 | 13 | 0 |

50.8% (n=124/244) of patients cared for in an escalation area experienced care in a non-clinical area such as a corridor, and 44.3% (n=55/124) of these patients were subsequently admitted.

### Hospital length of stay and 28-day all-cause mortality

The median hospital LOS for all admitted children was 2 days (IQR 1-3)Table 4. For those admitted having been cared for in an escalation area it was 2 days (IQR 1-3) and for those admitted without experiencing escalation area care it was 2 days (IQR 2-4). By 28 days 0.4% (3/815) of recruited patients had died, all having been admitted to hospital without having experienced escalation area care. See Table 4.

### Comparison between mixed and paediatric-only EDs

Of the recruited patients, either admitted or spending any time in an escalation area, 71.5% (n=583/815) were recruited from the 83 mixed EDs, with 28.4% (n= 232/815) of children recruited from the 13 paediatric-only EDs (Table 5).

**Table 5.**
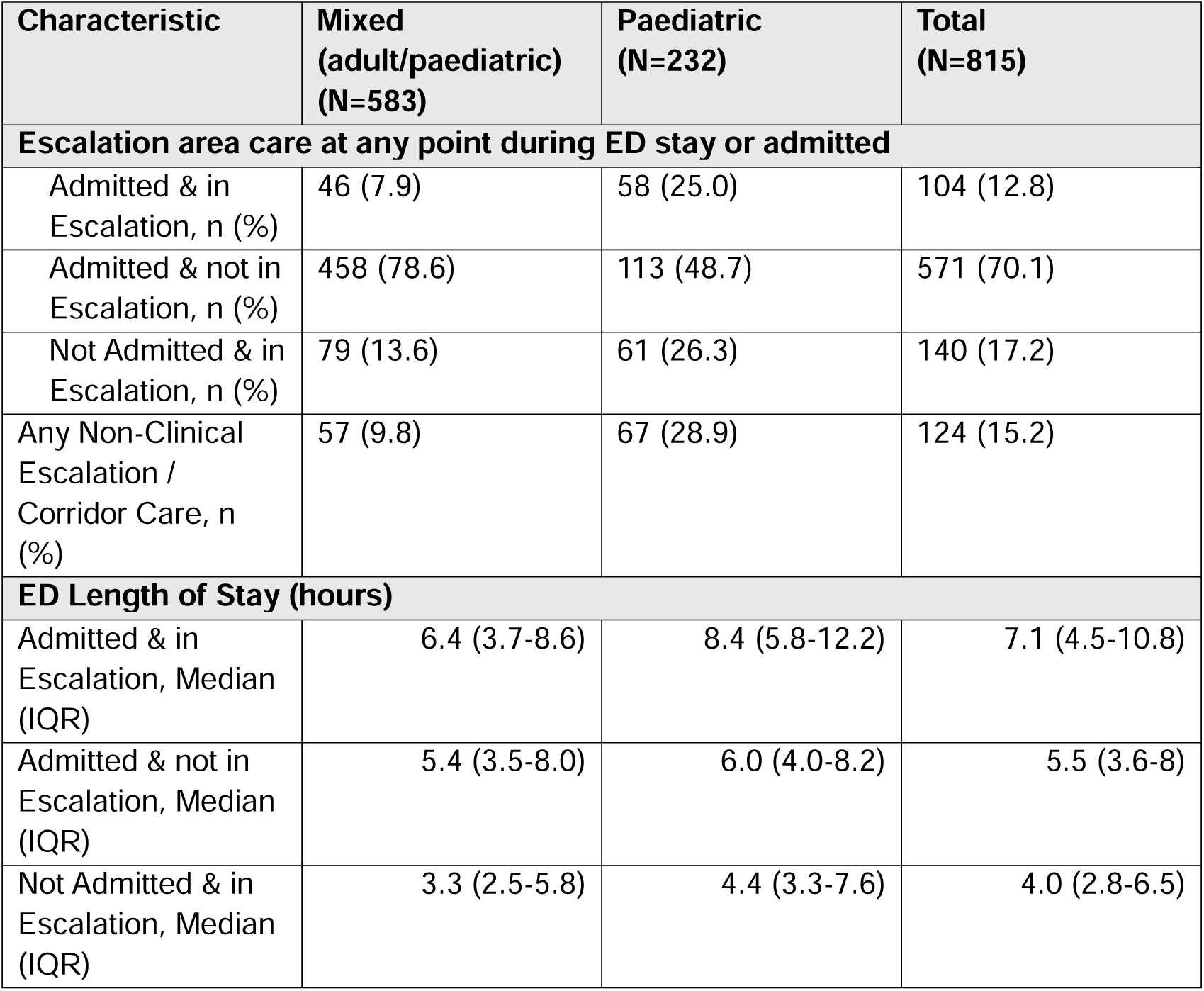
Escalation and corridor care experienced at any point during ED stay by hospital type.

Among those recruited in mixed EDs, 7.9% (n=46/583) were admitted having experienced escalation area care, 78.6% (n=458/583) were admitted without experiencing escalation area care and 13.6% (n=79/583) experienced escalation area care and were discharged. Among those recruited in paediatric-only EDs, 25.0%% (n=58/232) were admitted having experienced escalation area care, 48.7% (n=113/232) were admitted without experiencing escalation area care and 26.3% (n=61/232) experienced escalation area care and were discharged.

For those admitted having experienced escalation area care, ED LOS was 8.4 hours (IQR 5.8-12.2) in paediatric only EDs and 6.4 hours (IQR 3.7-8.6) in mixed EDs. Among those experiencing escalation area care at any point during their ED stay 56.3% (n=67/119) spent time in a non-clinical area such as a corridor in paediatric only sites with 45.6% (n=57/125) doing so in mixed sites.

## Discussion

### Main results

While most paediatric-only EDs did not use escalation areas during the study period, 6.6% (n=75/1,132) of children present in such EDs were seen to be receiving care in an escalation area during the snapshots. Among children admitted to hospital from mixed and paediatric only EDs: 15.4% (n=104/675) experienced escalation area care at any point during their ED stay with half of these experiencing care in a non-clinical area such as a corridor, median time spent in escalation areas was 2.73 hours (IQR 0.98-5.04). In EDs caring for adults and children, 1 in 10 (9.6%, n=8/83) sites reported caring for adults in paediatric areas. Among paediatric only EDs 69.2% (n=9/13) reported having no available resuscitation cubicle during at least one of the snapshots.

### Comparison to other studies

This analysis implies that the use of escalation area care in children is likely lower than in adults, where 17.7% of patients were seen to be experiencing such care at the time of the snapshots.^9^ However, this evidence demonstrates that contrary to both national RCEM guidance and RCPCH standards, escalation area care is widespread for children and crowding results in adults being cared for in paediatric areas.^7,8^ This is a clinical and safeguarding risk for children and places adult patients in an environment not designed for their clinical needs.^8^

Evidence from the US in 2011 and 2013, reported that for patients with asthma, ED crowding is associated with longer time to physician initial assessment and delays to corticosteroid administration.^10,11^ For paediatric patients with long bone fractures, crowding results in decreased quality of care, including administration of analgesia.^12^ This study saw half (50.8%) of the children admitted from the ED having experienced escalation care during their ED stay spending time in non-clinical spaces such as corridors, which are likely to have poor clinical monitoring. The median time spent in escalation areas of 2.73 hours (IQR 0.98-5.04 hours) is a significant patient safety concern, as children can deteriorate rapidly if significantly unwell.^13^ Evidence regarding the impact of crowding on patient outcomes in children is urgently required.

### Limitations

The limitations of the UNCORKED study methods are described in detail elsewhere and include the snapshot approach, lack of agreed definition for escalation area care and only clearly objective, visible criteria for identifying waiting room patients who should be in an ED cubicle.^8^ For this secondary analysis there are specific limitations to the interpretation of this data.

In those EDs seeing paediatric and adult patients a separate paediatric denominator was not reported. It was therefore not possible to provide an overall proportion of paediatric patients receiving escalation area care. A sub-group analysis of paediatric-only EDs is presented, however, it is possible that the escalation area care experience of these EDs is different to the situation in mixed EDs. This study has previously reported lower rates of escalation area care among adults in Major Trauma Centres compared to trauma units or local emergency hospitals, it may be the case that escalation area care occurs less frequently in specialised paediatric-only EDs comapred to mixed EDs hosted in trauma units or local hospitals.^9^

Length bias may be a consequence of the snapshot design where patients who have been in the ED for longer are more likely to be recruited to the study. The effect of such bias on this analysis is unknown; however, it is likely to have applied equally for each patient recruited. The snapshots were taken over a two-week period in March 2025 which may limit the generalisability of the findings to other periods of the year when the healthcare system may be under different levels of strain.

### Implications for Policy

The UK government is clear that corridor care is unacceptable and specifically highlights paediatric patients as a cohort who should never cared for in these areas. It should be a priority for policy makers to take urgent action to ensure this becomes a reality for paediatric patients and their families presenting to UK EDs.

### Conclusion

Escalation area care is a common and systemic feature of paediatric emergency medicine in the UK. It is also clear that the corridor care crisis is having a significant impact on children being cared for in mixed adult/paediatric EDs. This requires urgent action from ED clinicians, hospital management and healthcare policy makers to ensure future paediatric patients are cared for in appropriate ED environments and to ensure the crowding crisis in adult emergency medicine does not put children at any unnecessary safeguarding risk.

## Supporting information

Collaborators list

## Data Availability

All data produced in the present study are available upon reasonable request to the authors

## Notes

Funding: The study was funded by RCEM. RCEM grant number RCEM24_SG_4.

### Competing Interest Statement

The authors have declared no competing interest.

### Author Declarations

Ethical approval was gained from a Brighton and Sussex research ethics committee (ref: 24/LO/0837).

