## Supplementary material for "Paediatric escalation area and corridor care in the Emergency Department, a secondary analysis of the UNCORKED study": Collaborators list

Local collaborators: Aberdeen Royal Infirmary (Conal Mulholland, Toluwalase Fasina); Addenbrooke’s Hospital (Paul Bonhomme, Susie Hardwick, Asa French, Emma Clark, Jewel Inapan, Kerry Meynell); Aintree Hospital (Abdo Sattout, Richard Evans, Hayley Digby, Danielle Mclaughlan, Gemma Parker, Sarah Murphy); Airedale General Hospital (Martin Kelsey, Dan Raine, Charlotte Kelsey, Heather Collier, Virginia Clarke, Lindsey Stephens); Antrim Area Hospital (Kathryn Boyle, Jenny Sutherland, Matthew Copeland, Peter Devine); Barnet Hospital (Taranpreet Bhoday, Charlotte Walker-Jones, Jodie Mahendran); Barnsley Hospital (Fiona Dunning, Jenny Vernon, Scott Roantree, Emelia Barraclough, Hannah Brown); Basildon Hospital (Edward Lamuren, Mohamed Attia, Dawn Cairns, Shanais Thompson, Jane Thomas, Bushena Miyesa, Michael Villaruel, Princess Gabiana); Basingstoke and North Hampshire Hospital (Stian Mohrsen, Karolina Chalk, Tanja Border); Bedford Hospital (Devasena Subramanyam, Melchizedek Penacerrada, Jacinta Dias, Rachel Lorusso, Marielle Tolentino, Blessy Rethi, Christina Smith, Sushma Patil, Sanya Niyas); Bristol Royal Infirmary (Katherine Murdoch, Charlotte Munday, Katie Sweet, Ruth Bonsor, Linda Pipira, David Hopgood, Zoe Garland, Robert Eggen, Philip Anyelba Tankpara, Reeana Irani); Bronglais General Hospital (Mohamed Nasser, Subhasree Biswas, Heather McGuinness); Broomfield Hospital (Karen Rhodes, Stacey Cotterell, Rachael Arnold); Calderdale Royal Hospital (Huw Masson, Katherine Fulcher, Megan Walters, Emren Potinci, Rusha Saha, Ishak Rouf); Charing Cross Hospital (Ann Carroll, Claire Kelly, Daniel Martin, Lauren Nichols, Niamh Sargeant); Chelsea Chesterfield Royal Hospital (Nick Mani, Vittoria Sorice, Emma Moakes, Natasha Lee, Jonathan Spackman); Conquest Hospital (Leigh Greenland, Sarah Goodwin, Claire Rutherfurd, David Jones, Christy Biji, Timothy Kesington, Ghassan Youssef, Danielle Vidler, Peter Garner, Louise Watson, Janet Sinclair, Toni de Freitas, George Youssef, Oliver Reigler, Aparna Senjuti, Shabih Zahra, Kate Fever, Penny Boxall); County Staffordshire Hospital (Adebola Olorunfemi, Jennifer Thomas, Nenette Abano); Craigavon Area Hospital (Dervla McKenna, Mark Feenan); Croydon University Hospital (Mark McInerney, Salwa Adam, James Brown, Rahisha Maskey, Sruthi Sridhar, Thuta Swe, Sharan Thapa, Kushagra Gupta, Maryam Alhasan, Qusay Al-Zubaidi, Michael Masucci, Harman Bhandal, Victoria Estlin, Trushna Marpaka, Nadir Fadol, Christo Anto); Derriford Hospital (Daisy Carter, Rosalyn Squire); Dumfries Ealing Hospital (Chris Nordstrom, Mariam Sharafeldin, Rafhan Kazi, Ibrahim Ayvaz, Sheena Quaid, Oluwatofunmi Gbenedio, Cheryl Aiello); East Surrey Hospital (Csaba Szekeres, Edward Rippingale-Combes, Ellen Jessup-Dunton, Catherine Kloppenborg, Isaac Brookman, Abbas Kumail, Varshini Gaddameedi, Viktor Mordavszky, Emmeline Simpson, Yathin Thammaiah); Eastbourne District General Hospital (Leigh Greenland, Sarah Goodwin, Claire Rutherfurd, Kelly Booth, Kavitha Anoop, Paul Bailey, Budhaditya Sanyal, Asim Bela, Chloe Hall, Oliver Reigler, Aparna Senjuti, Shabih Zahra, Kate Fever, Penny Boxall); Epsom Hospital (Grace Blows, Lisa Evans, Rebecca Macfarlane); Fairfield General Hospital (Mark Richardson-Riley, Zoe Thomas, Carol Lunney, Monaza Saeed, Pamela Bradley, Rebecca Marie Gibson, Jijimol Anthony, Julie Newton); Frimley Park Hospital (Kyi Kyi Nwe, Noha Elgendy, Ainsley Reynolds, Teena Kunnath, Thomas Davies, Khaled Alshakaki, Aatif Butt); Glan Clwyd Hospital (Kenneth Igwe, Siva Seramanperuman, Rachel Manley, Annette Bolger, Bethan Roberts, Oluwatoyin Idowu); Glangwili General Hospital (Mohamed Nasser, Becky Icke, Charlotte Jones, Samantha Coetzee, Bethan Landeg, Bethan Morse-Browning); Glasgow Royal Infirmary (Joanna Quinn, Monica McKenna); Gloucestershire Royal Hospital (Taher Sharaf, Michael Connelly, Jen Griffiths, Teresa Tarling, Alicia Wailes, Carys Whitby, Michael Castillo, Jennie Lowdell, Kate Pinchassoff, Nick Vallotton, Ashleigh Pritchard); Great Western Hospital (Sian Thomas, Ayesha Mushtaq, Laura McCafferty, Tim Slade, Lauren Eady, Sahaj Romana, Alex Law, Urwa Chaudhry, Hannah Sennitt, Funmi Olagbaiye, Joy Egbiri, Will Peach, Alasdair Franks, Leanne Price, Hannah Glatzel, Rob Kirkham, Rejani Jayan, Funmilayo Fatile); Hillingdon Hospital (Hinal Patel, Katherine Lovejoy, Alaric Belmain, Prachi Avatade, Vandan Savani, Neha Rao, Vaibhav Todkari, Prudence Oliver, Jonathan Porritt, Natasha Mahabir, Latha Aravindan, Mariam Nasseri, Shweta Sharma, Ru Grinnell); Hinchingbrooke Hospital (Rehan Fareed, Gbemisola Jenfa, Hassan Khan, Mark Harvey, Jozen Obrique, Elena Marco-Illana, Rincy Kurian, Kemisola Ajide); Huddersfield Royal Infirmary (Huw Masson, Katie Fulcher, Iqra Sultan, Sangeeta Khadka, Havishma Sreedharala, Sadvika Padmanabhuni, Ishak Rouf, Yasoob Alaameri, Lily Boyle, Marium Akhtar, Temiloluwa Ijiwole, Toby Williams, Phyu Syn); Hull Royal Infirmary (Austin Smithies, Laura Caley, Phillipa Howell, Ana Ferreira); James Cook University Hospital (Owen Williams, Sian Dalgleish, Elizabeth Griffiths); James Paget University Hospital (Amir Guirguis, Helen Sutherland, Elva Wilhelmsen, Montana Boast); John Radcliffe Hospital (Charlotte Tickle, Alexis Espinosa, Abigail Harris, Tinelly Sambo, Rashadul Alam, Rebecca Dehavillande, Jasmine Harris, Ralph Houet, Rufino Magallano, Franceska Tchapdeu, Daniela Krouzkova, Helen Law, Dominique Georgiou, Clare Fitchett, Nga Man Law, Martina Iorio); Kettering General Hospital (Mohammed Elwan, Hannah Britton, Vishnu Sreelatha, Hathurusinghage Piyaratne, Sari Al Hajaj, William Madu, Chloe Stolarski, Harika Maddireddy, Salma Mohamed, Suryanarayana Pasupuleti, Khaled Soliman, Swarga George); King George Hospital (Darryl Wood, Faika Qazi, Kim Bovill, Sam King); Kings College Hospital (Edward Baker, Clare Finney, Abdur Faisal, Caitlin Spooner, Burt Vergara); Kings Mill Hospital (Lynne Allsop, Jill Woodhead, Jill Kirk, Cheryl Heeley, Philip Buckley); Kingston Hospital (Kay Philcox, Marian Di Vito, Anjana Mistry, Roshni Molls); Leeds General Infirmary (Najeeb Rahman, Sophie Griffin, Sasha De Prendergast, Charlotte Winder, Matthew Smith); Leighton Hospital (Sean Crossman, Natalie McCormack, Adewale Naiyeju, Mark Quiambao, Sherrymole Chettaniyil, Martin Griffin, Joanne Harold, Claire Gabriel, Richard Lowsby, Victoria Westwood); Lincoln County Hospital (Awais Ahmad, Kelly Hubbard, Katie Dorr, Catherine Wyatt, Sarah Shephardson); Luton Macclesfield District General Hospital (Matt Lynch, Alex Scott, Natalie Keenan, Hannah Brennan, Joanne Bradley-Potts, Jan Tomkinson, Rachel Smith, Megan Balmer); Maidstone Hospital (Ragavan Navaratnam, Rebecca Seaman, Laura Kent, Amy Ackerley, Mel Kelly, Maisie Quinney, Anna Tunnicliff, Corinne Selsby, Monica Bin Meh); Manchester Royal Infirmary (Thomas Bannister, Craig Ferguson, Charlotte Taylor, Richard Body, Rhea Saldanha, Mohamed Abouhemda, Rachael Quayle, Sabiha Akter, Qurat Ul Ain, Karolina Szarzanowicz, Dana Hegazy, Sreenath Duggi, Zainab Baye, Olabisi Temilayo Omotayo, Peter David, Fady Dahalan, Sana Nasir, Sian Baldry); Medway Maritime Hospital (Adebayo Da-Costa, Victor Anota, Louise Brassington, Perpetual Palmer, Morakinyo Fasakin, Oluwafemi Aina, Emmanuel Oduware, Linda Ofori, Patience Nkala, Mary Everett, Dilukshi Wickramasinghe, Suzanne Williams, Clarissa Madla, Mahdi Succar); Midland Metropolitan University Hospital (Ashley Dark, Manal Shakir, Chinnu Prince, Syeda Tamanna, Izuchukwu Nwolisa); Milton Keynes University Hospital (Shindo Francis, Young Chewe, Muni Akande, Oluwadamilare Adetuberu, Annith Jerry, Katy Canavan, Sneha Venaik, Wuraola Akande); Musgrove Park Hospital (Sarah Johnson, Emma Machin, Evelyn Owusu-Mireku, Chrissie Lawrence, Shauna Bartley, Steffi Jose); New Cross Hospital (Arvinth Soundarrajan, Emma Jenkinson, Githushan Gengaparam, Rose Hemmings, Naomi Brown, Olasimbo Akinbobola, Ross Evans, Pradeep Nagaraju); Ninewells Hospital (Ross Hendry); Norfolk and Norwich Hospital (Laura Lee); North Manchester General Hospital (Pedro Simoes, Bency Laiju, Karen Connolly, Helen T-Michael, Adele Fitzgerald, Robin Sebastian, Zahid Yusuf); North Middlesex University Hospital (David Sims, Kim Stallard, Jonathan Holliday, Nirmala Arulampalam, Amy Knowles, Amr Kash, Yousef Atta, Nitish Seeboruth); North Tees Hospital (Sophie Hindmarsh, Laura O’Rourke, Paula Correia, Amrutha Jinka, Hillie Corr); Northampton General Hospital (Aiden Pettet, Flora Gallamoza, Amna Khalid, Ethelwolda Goyena, Bincy Kariyadil, Maxine Foo); Northern General Hospital (Ashleigh Trimble, Alex Robertson, Charlotte Crapper, Helen Wanstall, Emily Dale, Anna Wilson, Jack Bardwell); Northumbria Specialist Emergency Care Hospital (Alex Russell, Mark Harrison, Hayley McKie, Tracy Smith, Anna Smith); Northwick Park Hospital (Chris Nordstrom, Oluwatofunmi Gbenedio, Tabassum Khan, Sheena Quaid, Parveen Kaur, Rafhan Kazi, Ibrahim Ayvaz, Mariam Sharafeldin, Andreea Cuciuc, Ikenna Ohanenye); Ormskirk Hospital (Chelcie Jewitt, Nyquist Mooteeram, Craig Rimmer, Moira Morrison); Peterborough City Hospital (Robert Lee, Christopher Edmunds, Natalie Temple, Shukurat Adeola Adebayo, Caitlin Back, Stephanie Bates, Helen Burtenshaw, Raquel Calcada, Alys Capell, Kerrie Cavanagh, Jayne Hanby, Trish Mazambani, Graeme McLintock, Reshma Ramachandran, Frincy Sijo, Lauren Woods, Ionela Sinanovic, Helen Wilson, Suzanne Woodhouse, Manali Lilani, Emily Smith, Tracey Peachey, Tessa Stoby, Lois Frome); Pilgrim Hospital (Awais Ahmad, Amy Faunt, Jane Upsall, Kimberley Netherton, Rajeshwar Ranganathan, Muhammad Mohsin Isar, Moustafa Abouelkheir); Pinderfields Hospital (Sarah Robertshaw, Sarah Buckley, Amy Major, Shamundeshwari Mathavan, Freddy Kheshwalla, Katherine Goff, Elina John, Mohammad Irfan); Poole Hospital (Charlotte Humphrey, Karen O’Toole, Emma Langridge, Yasmin de Ath, Lauren Bumpass, Aryn Azlan, Thankam Boniface, Esha Moghan, Ursula Gannon, Emily Doidge, Eliza Chwiecko); Princess Royal Hospital, Haywards Heath (Juliet Ariel, Hadis Reyhani, Carla Clegg, Victoria Elston, Lara McNeill, Ruchindra DeSilva, Chizoba Azie); Princess Royal Hospital, Telford (Adrian Marsh, Danyal Fiaz, Wael Fuhaid, Megan Brown, Jaseem Mohamed, Anthonia Maduabuchukwu, Sidharth Smithapushpan, Mohamed Musa, Ahmad Sultan, Joseph Paruchuru, Helen Diack); Queen Alexandra Hospital, Portsmouth (Christiane Vorwerk, Zoe Daly, Andrew Gribbin, Leah Sinclair, Blessing Otobhiale); Queen Elizabeth Hospital, King’s Lynn (Sharleen Siu, Hilary Thornton, Charles Nwankpa, Amrit Chaulagain, Hannah Greenacre, Ashish Kundu, Mohamed Imam, Yunusa Abdullahi, Joanna Rudnicka, Jemshad Koola Parambath, Sophy Shedwell); Queen Elizabeth Hospital, Woolwich (Lauren Matthews, Sharon Hall, Olivia Ballard, Reshma Kamar, Adheera Singh, Tasnuva Tamanna, Faisal Yousif, Devaan Dezylva, George Wilson, Arif Shareef, Semi Segbenu, Chloe Hicks, Arwa Shaikh, Samuel Booth, Aadil Farooq, Matt Burton, Khurrum Rasool, Ameena Sulaiman, Haris Rauf Mohammad); Queen Elizabeth the Queen Mother Hospital, Margate (Georgiana Scarlat, Hazel Ramos, Joanne Deery, Tracy Hazelton, Eva Beranova, Natalie Mercado); Queen Elizabeth University Hospital, Glasgow (Ryan McHenry); Queen’s Hospital Burton (Jamie Hunter, Alison Fletcher, Ainsley MacShannon, Mohsin Mirza); Queen’s Hospital, Romford (Darryl Wood, Jaspinder Kaur, Sam King); Rotherham Hospital (George Hamson, Simon McCormick, Rebecca Pugh, Rachael Gaffney, Rachel Walker); Royal Albert Edward Infirmary (Gemma Burrows, Amanda Ahmed, Emma Robinson, Sacha Connor); Royal Alexandra Hospital, Paisley (Jennifer Ross, Imogen Sandiford); Royal Berkshire Hospital (Thomas Rickaby, Georgia Coward, Ada Ukaulor, Romilly Gosling, Maria Nozdrina, Georgia Porter, Rana Awadalla, Anna Okada, Charlotte Knowles, Sheena Faloon, Liza Keating, Priyadharshini Ravi, Emel Yildirim, Myra Naeem, Nandini Tewari); Royal Cornwall Hospital (Rosemary Hartley, Muhammad Irshad, Sally Thomas, Adam Spencer, Georgia Moore, Oliver Vincent, Dan Bawden); Royal Derby Hospital (Jasmine Soo, Bashar Elwir, Alison Rockey, Lianne Hufton, Elisha Cousins, Andrew Tabner, Graham Johnson, Alison Fletcher, Ainsley Macshannon, Alison Matthews, Jamie Hunter, Mohsin Mirza, Bethany Harvey, Claire Fearn, Muamar Bayanan); Royal Devon and Exeter Hospital (Sam Scotcher, Ruth Webb, Harvey Thompson, Mahima Bhatt, Jasmine Collins, Thomas Christie); Royal Hampshire County Hospital (Stian Mohrsen, Tanja Border, Karolina Chalk); Royal Infirmary Edinburgh (Luisa Padovani, Ysabelle Thackray, Johanna Walter); Royal Liverpool Hospital (Wojciech Sawicki, Danielle Mclaughlan, Hayley Digby, Sarah Murphy, Gemma Parker, Christopher Speed, Harry Dale, Richard Evans, Jessica Hindley, Mary Brodsky, Allayna Doherty, Amy Doyle, Faustina Ravi, Jennifer Entwistle); Royal London Hospital (Fiona Mendes, Ivan Petkov Kisyov, Noemi Caponi, Fiona McMahon, Jake Hong, Michael Eason, Hannana Khatun, Emily Campbell, Nimca Omer, Fatma Mohammed, Hannah Stalker, Grace Tunesi); Royal Oldham Hospital (Hassaan Afzal, Louise Howard-Sandy, Angiy Vian-Michael, Jack Haslam); Royal Preston Hospital (Kirsty Challen, Jennifer Townsend, Kristal Dirisam); Royal Shrewsbury Hospital (Adrian Marsh, Helen Diack, Nabina Bhattarai, Sandi Angus, Alessandra Fraser-Pye, Nisha Pai, Mohammad Bhuiyan, Ikram Mohamoud, Aribah Naveed, Omer Mohamed, Roanna Craven, Mohamed Mobasher); Royal Stoke University Hospital (Adebola Olorunfemi, Nenette Abano, Ukraina Garcia, Sylwia Mowinska, Abira Aftab, Joanne Hiden); Royal Surrey County Hospital (Giorgina Blanco, Anna Vaulks, Sheila Mtuwa, Wuraola Adeoye, Diane Montoya, Kay Reynolds); Royal Sussex County Hospital (Sara Basha, Divya Rajaiah, Alyssa Jordan, Lovelin Regina, Gabrielle Alexander-Harvey); Royal United Hospital Bath (Emily Wilson, Lucy Howie, Genette Parsons, Katy Stevenson, Amanda King); Royal Victoria Hospital Belfast (Leanne Brown, Adeel Akhtar, Christopher Finnegan, Um e Roman Moughal, Peter Heaney, Erin Fisher, Joel Abraham); Royal Victoria Infirmary, Newcastle (Lucy Curtis-Holloway, Chris Wilkinson, Elizabeth Clayton, Jonathan Bennett); Salford Royal Hospital (Dominic Kay, Ahmed Ali, Reece Doonan, Jessica Pendlebury, Stephanie Lee, Lisa Swindells, Sharon Barber, Jacob Hadfield); Salisbury District Hospital (Jenna Plank, Sophia Strong-Sheldrake, Abby Rand, Frederick Gleadowe, Emily Brockbank, Rory Catton, Matthew Charlwood, Fenia Dandelou); Scarborough General Hospital (Ed Smith, Francesca Duncan, Alison Turnbull, Laura Barman, Rachel Harrison, Dana Groom, Anna Waine, Rachel Harrison, Sakshi Beotra); Scunthorpe General Hospital (Kelum Perera, Sue Spencer, Joanne Hill); Southampton General Hospital (Caroline Thomas, Sidra Jamil, Elizabeth Frost, Anna Foster, Rachel Schranz, Abigail Johnston, Kate Sheppard, Matt Morris, Owen Gregory, Ines Moreira, Kerry Thorpe, Iberedem Umana, Kyla Brand); Southend University Hospital (Joanne Galliford, Sharon Tysoe, Anne McPherson, Eugene Mphansi, Lesley Nichols, Swapna Kunhunny, Abena Tweneboah, Oluwaseyifunmi Juba); Southmead Hospital (Chris Goodwin, Michael Gammon, Joshua Evans, Mohamed Ali, Tanaz Padiyath, Alice Ostojic, Will Sharp, Tarn Stroud, Aimee Wilkinson, Alex Sedgley, Naina Mistry, Olivia Sinclair, Harriet Jones, Helen Emery, Sandeep Nair, Emma Godson, Aran Jamieson, Fraser Birse); Southport Hospital (Craig Rimmer, Nyquist Mooteeram, Lorraine Bickerstaffe, Sabina Koprowska, Moira Morrison, Kerri Bowness, Chelcie Jewitt); St George’s Hospital (Jared Charlton-Webb, Phil Moss, Laila Altaffi, Dariush Micallef, Annalie Lahoud, Rawan Mouhandes, Nisha Gurung, Thomas Hosfield, Cayla Marshall, Shaimaa Mohamed, Muhammad Faham Saleem, Rosie Wright); St Helier Hospital (Grace Blows, Lisa Evans, Melissa Hanger, Hannah Brotherwood, Rebecca Macfarlane); St James’s University Hospital (Najeeb Rahman, Manou Sundararaj, Sasha De Prendergast, Charlotte Winder, Matt Smith, Katie Nolan, Sophie Griffin); St John’s Hospital (Johanna Walter, Ysabelle Thackray, Luisa Padovani); St Mary’s Hospital (Michael O’Connor, Ruud Nijman, Purushotham Harsha Vardhan, Saranya Ravindran); St Peter’s Hospital (Santosh Pradhan, Lok Thapa, Ahmad Saqer, Amit Bhandari, Donna Edano); St Richard’s Hospital (Andrew Pantelides, Clemence Lagroy de Croutte, Abbie Tutt, Tessa Mitchell, Usama Hassan, Siraj Mohammed, Rupali Sachdev); St Thomas’ Hospital (Laura Hunter, Ella MacInnes, Amy Harris, Miranda Smith); Stepping Hill Hospital (Gopala Pureti, Julie Melville, Diane Daniel, Chioma Akinfenwa); Sunderland Royal Hospital (Phil Dowson, Michael Thompson, Kyna Richardson); Tameside General Hospital (Mohammad Islam, Hussain Ahmad, Christy Simon, Roxanne Gray); The Cumberland Infirmary (David Miller, Jane Gregory, Theresa Cooper); The Grange University Hospital (Mary Kivell, Alastair Richards, Shiney Cherian, Claire Price); The Princess Alexandra Hospital, Harlow (Roberta Branisteanu, Daniel Hodges, Patricia Nabayego, Amara Benson, Sylwia Goliaszewska, Bibi Badal, Samantha Beck, Donna Foster, Louise Barnard, Joanne Finn); The Royal Bournemouth Hospital (Charlotte Humphrey, Annamaria Wilce, Javen Ramsami, Eliza Chwiecko, Declan Woodhouse, Danier Parker, Keerthana Aravindhan, Angela Healey, Lorenzo Bianchi, Helena Dunn, Omar Elsobky, Farzana Karim); Torbay Hospital (Elizabeth Florey, Joan Redome, James Allen); Tunbridge Wells Hospital (Ragavan Navaratnam, Rebecca Seaman, Laura Kent, Amy Ackerley, Mel Kelly, Maisie Quinney, Anna Tunnicliff, Corinne Selsby, Monica Bin Meh); Ulster Hospital (Andrew Dobbin, Jonny Taylor, Ellen Hirst, Vicki Adell, Rachael Johnston, Sara Molloy, Eleanor Flynn, Jonathan MacCorkell, Aine Forester, Jennifer Holmes, Caoimhe Sheppard, Emma McCann, Sarah Moorhead, Lauren Brown, Jill McGregor, David McKinney); University College London Hospital (Mohamed Abdalla, Samer Elkhodaor, Erlyn Lomibao, Bobby Garcia, Fabiola Sevilla Perez, Christopher Griffiths); University Hospital Coventry (Llinos Hutchings, Jordan Simms, Rachel Rose, Margaret Lindsay, Ibrahim Abdelrhman, Louise Bromwich, Karys Noonan, Adel Bilal, Mehmet Yilmaz, Caroline Leech); University Hospital Crosshouse (Jessica Jameson, Struan Powrie); University Hospital Hairmyres (Hayley-Isabella Cawley, Maria McLaughlin, Emma Speake, Keir Brown, Heather Liddell, Elliot Craven, Linzi Marie Clark); University Hospital Lewisham (Anna Colclough, Joemar De La Pena, Easteen Davis-McIntosh, Neisha Rhule, Blessing Kazooba, Liban Bussuri, Daniel Beasley, Nicholas Wilson, Adam Durbin); University Hospital Monklands (Nicola Moultrie, Fiona Hunter, Roisin McGovern, Tracy Baird); University Hospital of North Durham (Lauren Ferguson, Iain Fraser, Jamie Greenwood); University Hospital of Wales (Peter Tytler, Nicholas Manville, Rhys Thomas, Nathaniel Williams, Mark Crothers); University Hospital Wishaw (Chris Moultrie, Claire Beith, Karen Black); Victoria Hospital Kirkcaldy (Naomi Gunn, Chloe Haigh, Jacqueline James); Walsall Manor Hospital (Misbah Mohammad, Midhat Tahir, Ben Jones, Rachel Pearse, Rhianne Grice, Andre Fernandes); Warwick Hospital (Rachel Dancer, Johannes Du Toit, Penny Parsons, Judy Shirley, Camilla Stagg, Maxine Turner, Angela Day, Indy Atwal, Pamela Berry, Bolanle Ekisola, Bridget Campbell); Watford General Hospital (Daniela Burlacu, Arin Bose, Alice Balaican, Marietta Pilarska, Tanvi Patel, Jasmin Pandhal); West Cumberland Hospital (David Miller, Rosemary Harper); Weston General Hospital (Hazem Amer, Deepika Mittal, Susan Wilkinson, Mandie Williams, Catherine White, Edel Robbins, Katrina Stallard, Janet Parker, Joshua Christie); Wexham Park Hospital (Nicolas Marquinez Vecchione, Sarah Wilson, Anrhona Galloway, Neeraja Mini Mol, Rhoda Law-Onilearo, Molly Everett, Christine Del Rosario, Benjamin Rush, Karen Chivers, Annie Green, Isha Chhetri, Eleanor Hassett, Wameedh Almaalullah, Mohamed Badawi, Maria Cruz, Chaw Myo, Hritik Nautiyal, Nicole Kader, Joana Da Rocha, Heather Bonner); Whipps Cross University Hospital (Fiona Mendes, Igho Jude Mukoro, Jack T G P Maloney, Mohamed Elymany, Michael Tadres, Alan Padayathil, Saloni Phor, Muhammad Farooq Mamoon); Whiston Hospital (Oliver Moore, Harriet Pleasant, Sharon Burnett, Jennifer Procter, Sharon Dealing, Jayne Evans, Kerri Bowness, Zoe Grindley, Eric Mbogu, Anamika Algeo, Karen Shuker, Robert Fuller); Whittington Hospital (Rachel Johnston, Leanne Taylor, Rahman Ahmad); William Harvey Hospital (Alison Brown, Reanne Solly, Olajumoke Owolabi); Withybush General Hospital (Mohamed Nasser, Michelle Edwards, Kelly Wood, Catherine MacPhee); Worthing Hospital (Laurence Caines, Isabel Norris, Kirsten King, Mohamed Selim, Andrew Thomson, Shahnaz Hakeem, Syed Muzaffar Hasan Kirmani, Alexander Dalton, Lorena Lucioli, Muhammad Sulaman Ashraf, Hannah Lidbetter); Wrexham Maelor Hospital (Ash Basu, Rachel Hughes, Rebecca Pope, Aimi Streeter, Phil Metcalf, Pavan Mangalore, Rob Fenwick, Nina Bassett, Jomcy John, Sarah Garrett); Yeovil Hospital (Stevan Bruijns, Rebecca Covey, Lucy Pippard, Nigel Beer); York District Hospital (Deborah Goldfield, Catriona Laverty, Katherine Atley, Stephen Grace, Christopher Bourlet, Laura Wendon; Ysbyty Gwynedd (Bangor) (Georgina Keyte, Catrin Davies, Ateev Juneja, Jessica Flint, Felipe Pellizon, Jess Trevett, Delyth Davies, Wendy Scrase).
